# Improving cardiovascular population risk in primary care: protocol for the PROSPERA cluster-randomized controlled trial of a complex multilevel intervention

**DOI:** 10.64898/2026.02.13.26345729

**Authors:** Vera A. M. C. Bongaerts, Rimke C. Vos, Steven H. J. Hageman, William K. Redekop, Jannick A. N. Dorresteijn, Mattijs E. Numans, Hendrikus J. A. van Os

## Abstract

**Introduction:** The growing burden of cardiovascular disease (CVD) threatens the sustainability of our healthcare systems. In the Netherlands, two-thirds of CVD-related healthcare is delivered in primary care practices, primarily by practice nurses. Given the increasing staffing shortages and the substantial heterogeneity of the population of primary care patients at increased risk of CVD, cardiovascular risk management strategies should be better tailored to individual patients’ needs and risk profiles. Therefore, we developed the PROSPERA program, aimed at the population of patients enrolled in integrated care programs for cardiovascular risk management in primary care.

**Methods and analysis:** The PROSPERA study is a pragmatic 1:1 cluster randomized controlled trial in 22 primary care practices in the greater Leiden and the Hague region in The Netherlands, investigating real-world effectiveness of the PROSPERA program with care as usual. The PROSPERA program is a multilevel complex intervention, including a population-level risk stratification and three individual patient-level components: a healthcare professional training program in cardiovascular risk communication, a lifestyle questionnaire (Lifestylecheck), and a clinical decision support tool (U-Prevent). Patients between 40–90 years of age enrolled in integrated care programs for increased cardiovascular risk are included. The primary outcome is defined as successful achievement of all protocol-defined cardiovascular preventive treatment goals, measured as the difference between the proportion of patients who meet the outcome in the intervention versus the control group at 18 months. Secondary outcomes include implementation outcomes, healthcare professionals’ satisfaction on usability of the PROSPERA program, patient experience with shared decision making and decisional conflict, and outcomes on cost-effectiveness.

**Ethics and dissemination:** Consent for the use of routine healthcare data is obtained through an opt-out procedure for participating practices. Digital informed consent will be obtained for the study questionnaires for patients and healthcare professionals. Main findings on effectivity, efficacy and implementation will be disseminated via peer-reviewed journals and via (inter)national conferences.

**Trial registration number:** NCT06593704

**Strengths and limitations of this study:**

- The PROSPERA program was co-developed with stakeholders and refined via a preceding qualitative study, ensuring better integration with clinical workflows.
- The pragmatic design of this study allows for broad inclusion at the level of the population at increased risk of CVD, enhancing representativeness and inclusivity.
- Use of the Extramural Leiden Medical Center Academic Network (ELAN) data infrastructure enables comprehensive collection of healthcare data.
- The complexity of the current information- and technology (IT) infrastructure prototype is a constraint, but is expected to be simplified in the future.
- Potential cross-over contamination cannot be prevented, as control practices may have informal access to some PROSPERA components, although these are not yet widely implemented.

## INTRODUCTION

Cardiovascular diseases (CVD) are the leading cause of death for people worldwide (1). The growing disease burden and the associated costs endanger the sustainability of our healthcare systems (2). Therefore, improving prevention of CVD is key, since most of the risk is caused by modifiable risk factors (3). In clinical practice, cardiovascular risk management (CVRM) is usually addressed at the level of the individual but is generally insufficient at a population level. This is supported by the wide variation in percentages of CVD risk factor control across the cardiovascular population (4-6).

In the Netherlands, about two-thirds of the patients at risk of CVD are treated in primary care practices by a general practitioner (GP) or a practice nurse (PN) in integrated care programs (4). These are programs developed to ensure a continuum of care for patients with chronic conditions such as increased risk of CVD or diabetes mellitus (DM) – a population with substantial heterogeneity regarding individual cardiovascular risk and patient needs (7). However, the base frequency of CVD consultations in integrated care programs is currently not determined by the magnitude of a patients’ cardiovascular risk, and there is at present no structure in place to identify patients for prioritized outreach. Hence, present CVD integrated care programs can largely be considered a ‘one-size-fits-all’ approach, while tailoring cardiovascular prevention to the needs of the individual patient may be crucial to ensure CVRM stays accessible and in compliance with current guidelines (8, 9).

In recent years, advances in availability of routine care data and cardiovascular risk prediction models have enabled automated individual risk prediction. This has led to development of clinical decision support tools for individual risk predictions (10). However, for healthcare professionals in primary care dealing with increasing prevalence of patients at risk of CVD and growing staffing shortages, the use of clinical decision support tools in the consultation room is still suboptimal (11, 12). Taking this into account, a logical next step would be to pursue a multilevel approach that improves CVRM integrated care programs on (i) a population level by using a proactive and risk-based approach for prioritised patient outreach, and (ii) optimise prevention at the individual patient level by better integrating individual clinical decision support tools for cardiovascular risk prediction that support shared decision making in the consultation room (13). This multilevel approach would allow for prioritising preventive care strategies to those who most need it, while simultaneously complying with guidelines and other specific needs of the individual patient.

Therefore, we developed the PROSPERA program, a multilevel complex intervention addressing both the population level and the level of the individual patient (14). At a population level it provides the healthcare professional with the opportunity to proactively direct available healthcare resources based on differing risk levels in the CVD population, differentiating between who is most in need of intensive treatment versus who may require less frequent consultations. At the individual patient level, the PROSPERA program consists of (i) a healthcare professional training program in cardiovascular risk communication, (ii) an individual-level Lifestylecheck questionnaire, and (iii) an individual-level clinical decision support tool (U-Prevent).

The pragmatic PROSPERA cluster Randomized controlled trial (cRCT) follows a preceding qualitative study (14, 15), and aims to improve the cardiovascular risk profile for the population of patients enrolled in integrated care programs for increased cardiovascular risk in primary care. Secondary outcomes include implementation outcomes (fidelity and penetration), healthcare professionals’ satisfaction on usability of the PROSPERA program, patient experience on shared decision making and decisional conflict, and cost-effectiveness.

## METHODS AND ANALYSIS

This study protocol is reported in accordance with the *Standard Protocol Items: Recommendations for Intervention Trials* (SPIRIT) reporting guidelines.

### Study design

The PROSPERA cRCT follows a pragmatic hybrid type I implementation- and effectiveness design to address both effectiveness and implementation outcomes within one study (16). The PROSPERA cRCT will be conducted in primary care practices affiliated with the Extramural Leiden University Medical Center Academic Network (ELAN) in the greater Leiden and the Hague region, the Netherlands, containing approximately 150 primary care practices. Figure 1 shows an anticipated randomization infographic. Clustered randomization to either the intervention (PROSPERA program) or control group (care-as-usual) will be performed in a 1:1 ratio on the level of primary care practices, through Castor Electronic Data Capture via a validated variable block randomization model (17). The anticipated number of eligible patients will depend on the size of the participating primary care practices. Allocation will only be visible for the researcher after inclusion and randomization of the primary care practice. Blinding of primary healthcare professionals and patients will not be possible due to the nature of the intervention. The study is expected to span a total duration of 18 months. Figure 5 shows a schematic representation of the timeline.

**Figure 1.**
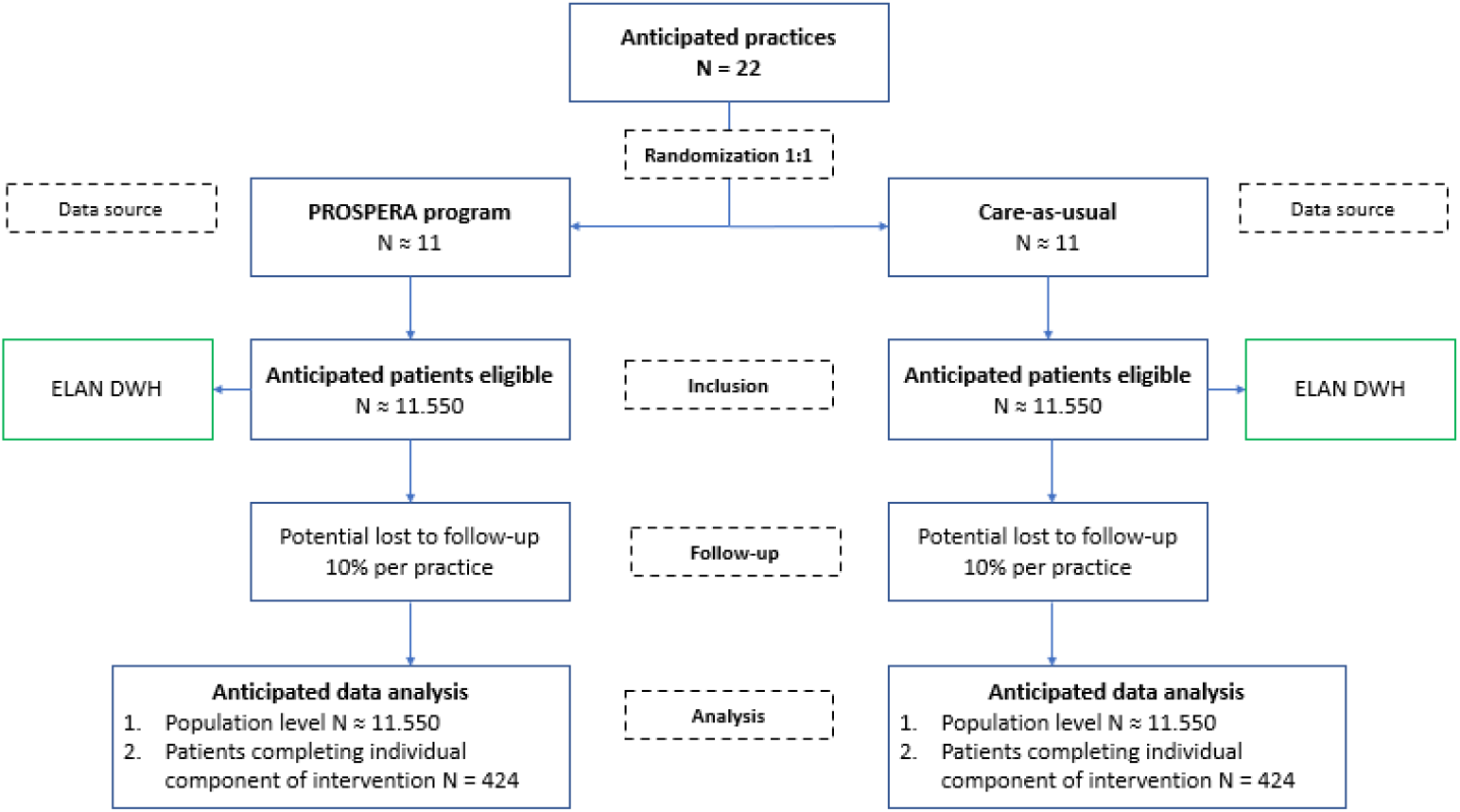
Anticipated randomization infographic *\*ELAN DWH = Extramural Leiden University Medical Center Academic Network Data Warehouse*.

### Study population

Patients are eligible for inclusion if they meet all the following criteria: age ≥ 40 years and ≤ 90 years, enlisted with one of the participating primary care practices and enrolled in one of the Dutch integrated care programs for (increased risk of) CVD or DM. Patients will be excluded if they receive palliative care, are pregnant, are pre-dialysis or in a kidney transplantation procedure, have a form of cognitive impairment, are treated in other integrated care programs because of their frailty (e.g. elderly who are treated in the ‘Frail Elderly integrated care program) or if they are diagnosed with diabetes mellitus type I.

### Care-as-usual

The comparator in this study will be standard cardiovascular risk management provided by primary care healthcare professionals in integrated care programs for (increased risk of) CVD or DM. This care-as-usual means that patients with an increased risk for cardiovascular disease receive annual invitations for CVRM consultations to ensure continuity of care in accordance with the Dutch Cardiovascular Risk Management Guidelines of General Practitioners (9, 18). Within these programs, the GP – and not the specialist in the hospital – must provide CVRM, and patient motivation is required for enrolment. Practice nurses are often the professionals that execute the consultations within the integrated care programs.

### Intervention

The PROSPERA program constitutes a complex intervention, and its development, implementation and evaluation will be conducted according to the United Kingdom Medical Research Council (MRC) framework for developing and evaluating complex interventions and follows the four phases for complex intervention research: (i) development of the intervention ii) feasibility testing iii) evaluation and iv) implementation (14). For this reason, the development and feasibility of the PROSPERA program and its accompanying IT structures were tested in a qualitative study before initiating this subsequent cRCT (15, 19, 20). As a result of insights gained in this preceding qualitative study, minor adaptations were made to the PROSPERA program and supporting implementation strategies were put into practice.

Figure 2 shows a schematic representation of the intervention components of the PROSPERA program. The PROSPERA program consists of interconnected components designed to deliver proactive and targeted CVRM. The process flow for the healthcare professional is as follows: the program will start with a risk stratification on population level for eligible patients and prioritized outreach. On the individual patient level, it follows with an accredited training for healthcare professionals to improve cardiovascular risk communication in the consultation room. Finally, the program continues with personal prevention for the individual patient using U-Prevent and the Lifestylecheck during consultation (10, 21).

**Figure 2.**
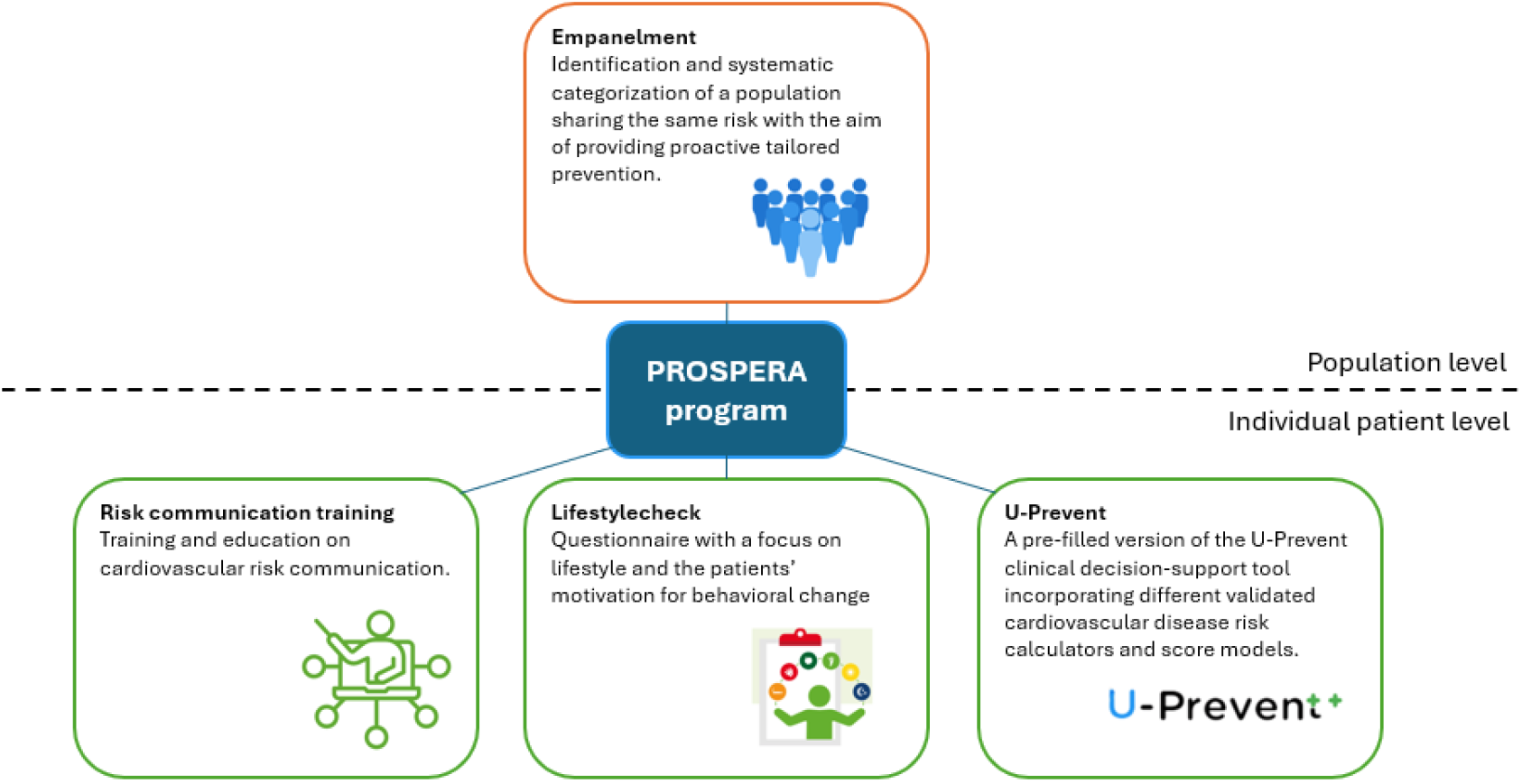
Components of the PROSPERA program

#### Empanelment

The population level component of the PROSPERA program comprises an empanelment approach. Figure 3 shows a schematic representation. Empanelment involves risk stratification of the population and provides healthcare professionals with the opportunity to proactively select patients for CVRM. Creation of the PROSPERA panels is based on two criteria. First, whether the patient requires primary (apparently healthy) or secondary (history of CVD) cardiovascular prevention. Second, whether the patient has achieved the pre-determined preventive CVD treatment goals based on the European Society of Cardiology (ESC) Clinical Practical Guidelines, the Dutch Cardiovascular Risk Management Guidelines of General Practitioners and the Dutch Diabetes Mellitus type 2 Guidelines of General Practitioners (8, 9, 22). This leads to three PROSPERA panels: normal, high, or extra-high. Further details on the panels and treatment goals are outlined in Appendix A. Panels are updated every three months. The first empanelment marks the starting point of the PROSPERA cRCT (T=0).

**Figure 3.**
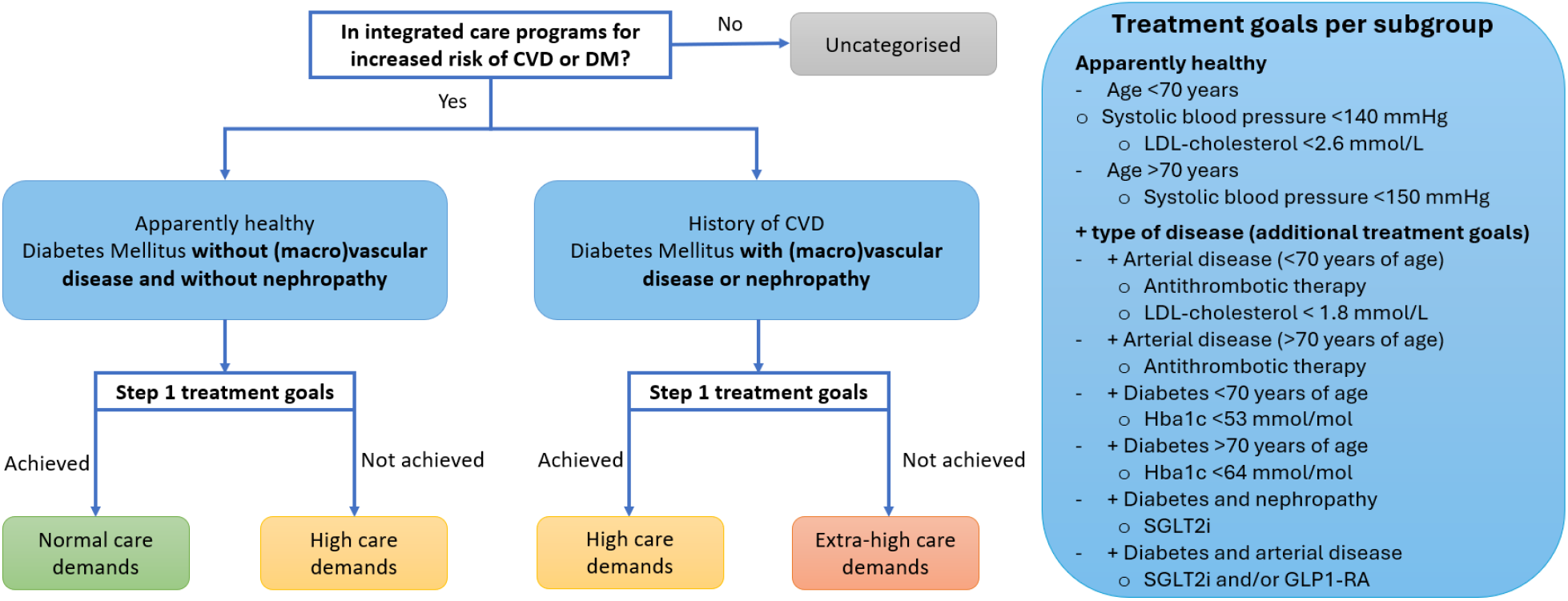
Empanelment and protocol defined treatment goals per subgroup

Missing clinical data for patients in integrated care programs, such as missing blood pressure measurements, missing LDL-cholesterol levels or missing HbA1c levels for DM patients, that are missing for >2 years, will be considered as treatment goals not achieved, given that individuals enrolled in integrated care programs are invited on a regular basis for risk factor control.

Healthcare professionals are informed about how the panels are created and are then provided with a panel list by the researcher. The panel list provides a basis for prioritized outreach, complementing the existing knowledge of healthcare professionals regarding their population at risk of CVD. Healthcare professionals may exercise their professional judgment in determining how to use this list for prioritization. Patients who are invited continue with the Lifestylecheck and U-Prevent components of the PROSPERA program during consultation.

#### Risk communication training

Healthcare professionals will attend a mandatory accredited training session on cardiovascular risk communication. Learning objectives include the translation of cardiovascular risk- and benefit calculations from U-Prevent to understandable information for the patient. The training will cover topics like the concept of lifetime risk, 10-year risk, CVD-free life expectancy, and number needed to treat (NNT). Healthcare professionals will be provided with the opportunity to review the recorded training online and attend question-and-answer sessions with the trainers.

#### Lifestylecheck

The Lifestylecheck is a questionnaire dedicated to promoting healthier lifestyle behaviours in the Dutch population (21). In the PROSPERA program the Lifestylecheck will be adopted as a pragmatic tool to discuss current lifestyle and behavioural change incentives for eligible patients with an increased risk of CVD. The patient is asked to complete the questionnaire as preparation before consultation. Since patients indicate their motivation for behavioural change on the questionnaire, it can be used by the healthcare professional to guide motivated shared decision making in combination with the U-Prevent clinical decision support tool (21, 23).

#### U-Prevent

The U-Prevent medical device is used as a clinical decision support tool during consultation between healthcare professional and patient. U-Prevent incorporates different validated CVD risk calculators and score models in one digital application (10, 24, 25). Score models used in U-Prevent to predict 10-year cardiovascular risk are SCORE2, SCORE2-OP, SCORE2-Diabetes and SMART2. The SMART-REACH, DIAL2 and LIFE-CVD2 score models are used for lifetime risk and treatment-modelling predictions. The device complies with the transitional requirements of the Medical Device Regulation (EU 2017/745). The use of U-Prevent is endorsed in Dutch General Practice Standards for Cardiovascular Risk Management and U-Prevent is currently used on a small scale by healthcare professionals working in both primary and secondary care in the Netherlands (9, 18, 24). The version of U-Prevent used in this study is adapted for study purposes by pre-filling available relevant risk predictors such as biographical information, clinical measurements, laboratory findings and current medication use from the electronic health records to ease the workflow of the healthcare professional.

### Data management and infrastructure

Data will be obtained via questionnaires and via ELAN. ELAN is a dynamic data infrastructure originating from clinical-academic collaboration and supported by quarterly extracted pseudonymized routine healthcare data from affiliated organizations such as hospitals and primary care practices (26). Data from ELAN is only accessible upon request and with permission from the ELAN committee. In the PROSPERA study this routine healthcare data will be used for the empanelment, the pre-filling of U-Prevent and for assessing the primary and secondary outcomes. It includes, amongst others, coded clinical measurements, laboratory findings, information on medication use and information on smoking behaviour (26, 27). Identifiable patient data are pseudonymized for researchers and other stakeholders, but accessible for healthcare professionals via a trusted third party in the Netherlands responsible for data processing (see figure 4) (28). A Data Management Plan is in place at dmponline.lumc.nl and accessible upon request.

**Figure 4.**
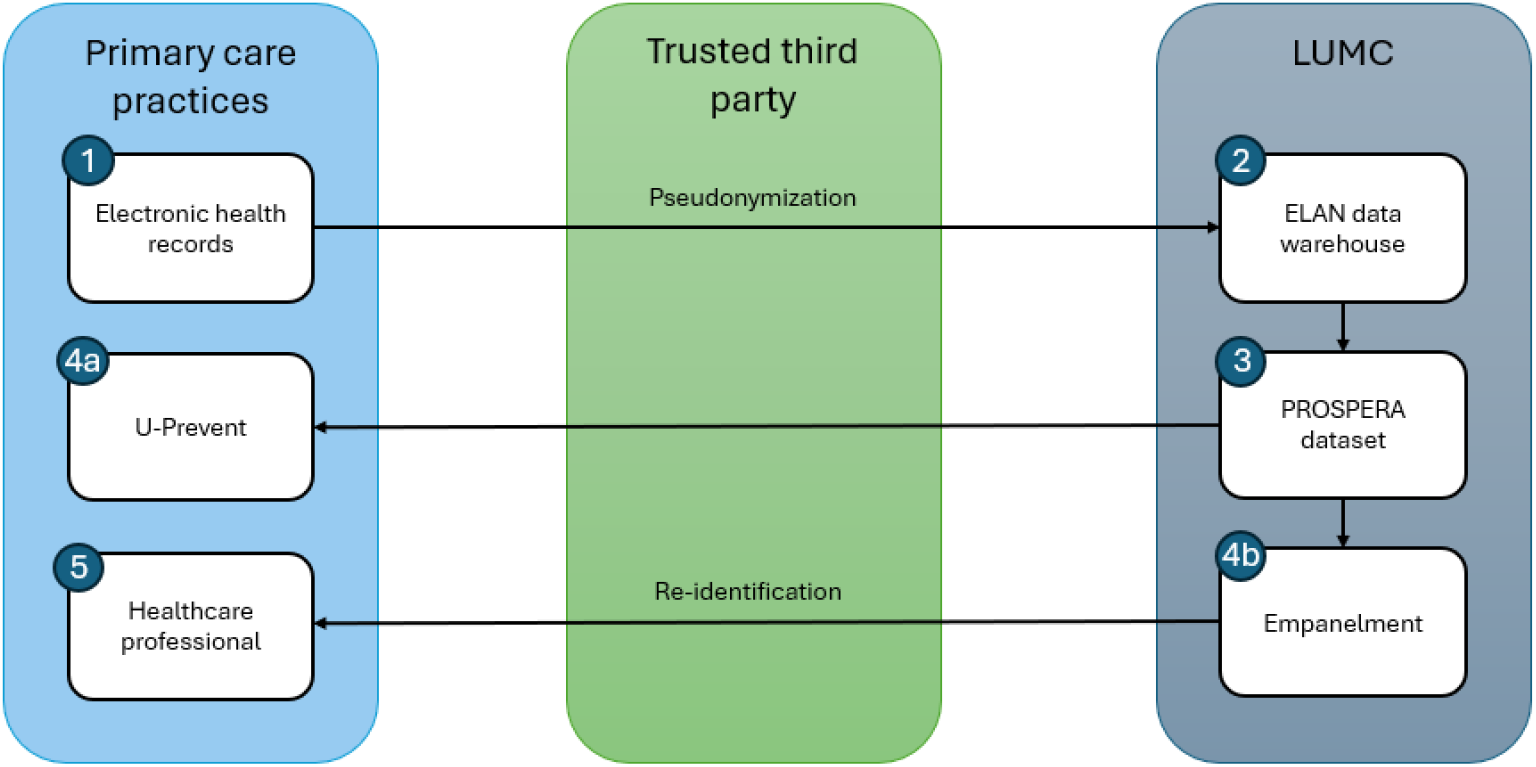
PROSPERA data infrastructure Steps: 1. Routine healthcare data are extracted from electronic health records in primary care practices. Data are pseudonymized via a trusted third party. 2. Data are stored in ELAN data warehouse. 3. Data are requested for research. 4a. Data are used to pre-fill in U-Prevent. 4b. Data are used for PROSPERA empanelment. 5. Healthcare professionals have access to re-identifiable data to be used in clinical practice.

**Figure 5.**
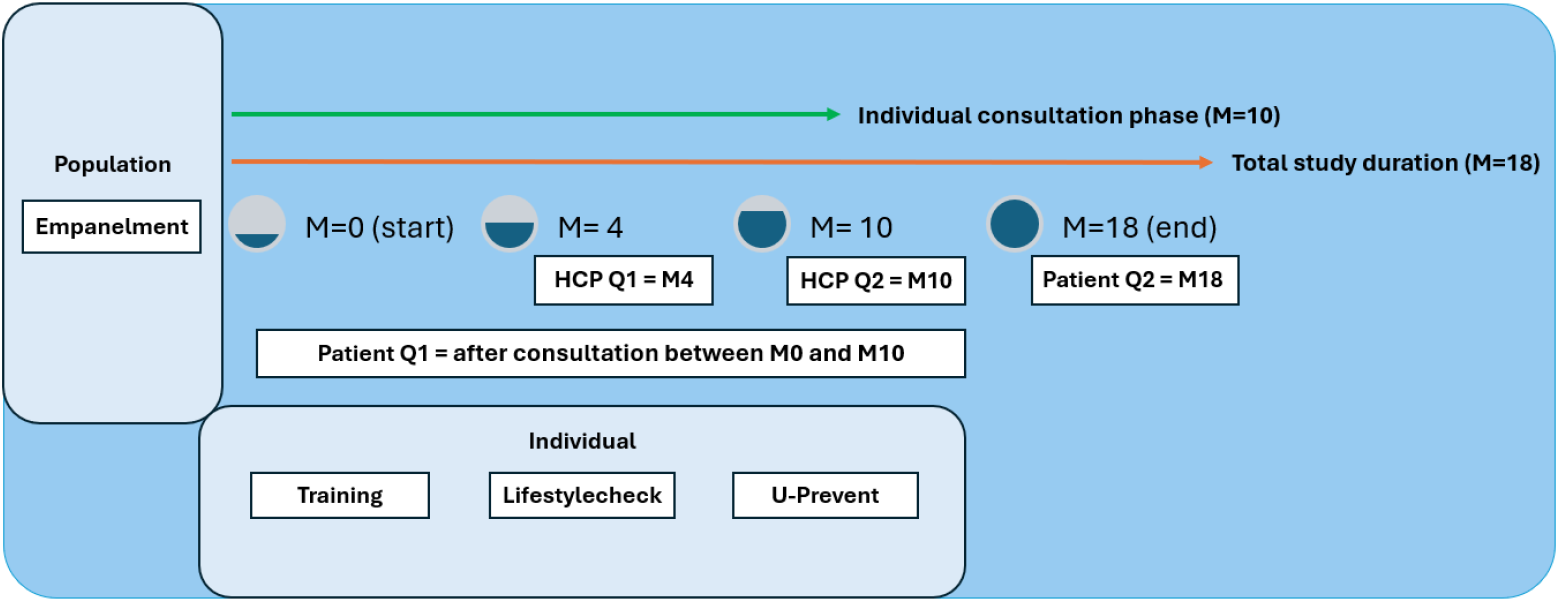
Timeline of study procedures for healthcare professional (HCP) and patient per month (M)

### Primary outcome

The primary outcome is defined as successful achievement of all protocol-defined cardiovascular preventive treatment goals, measured as the difference between the proportion of patients who meet the outcome in the intervention versus the control group at 18 months.

### Secondary outcomes

Secondary outcomes include the implementation measures fidelity and penetration, outcomes on healthcare professionals’ satisfaction on usability of the PROSPERA program, outcomes on patient experience with shared decision making and decisional conflict, and outcomes on cost-effectiveness. Secondary patient outcomes will be assessed in a subgroup only. Table 1 shows the specifications per questionnaire.

**Table 1.** Specifications per questionnaire.

| Patient | Questionnaire | Measurement moment (MM) and time duration | Number of items | Range | Interpretation |
| --- | --- | --- | --- | --- | --- |
| Cost effectiveness | EQ-5D-5L | MM = after the intervention and after 12 months. Time: 2 min. | 6 | 5-point scale (converted to a 0-100 range) | Low scores indicate the best health state, high scores indicate health problems |
| Shared decision Making | Shared Decision-Making Questionnaire (SDMQ-9) | MM = after the intervention. Time: 3 min. | 9 | 6-point scale (converted to a 0-100 scale) | Higher scores indicate a greater extent of shared decision making |
| Decisional conflict | Decisional Conflict Scale* | MM = after the intervention. Time: 3 min. | 16 | 3-point scale (converted to a 0-100 range) | Higher scores indicate more decisional conflict |
| HCP | Questionnaire(s) | Estimated time | Number of items | Range | Interpretation |
| Fidelity | PROSPERA Fidelity Scale | MM = at month 4 and 10. Time: 5 min. | 16 | 0-100 scale | Higher scores indicate higher fidelity and adherence |
| Usability | System Usability Scale (SUS) | MM = at month 10. Time: 2 min. | 10 | 5-point scale (converted to a 0-100 range) | Higher scores indicate higher usability |

\*The low literacy version translated to Dutch.

#### Implementation outcomes

Fidelity is the extent to which the PROSPERA program components are used as intended. Fidelity will be assessed in two ways: 1) through a PROSPERA Fidelity Scale for primary healthcare professionals designed for the purpose of this study (see appendix B), and 2) as the percentage of the target patient population that has received the U-Prevent component of the PROSPERA program during the study duration, as measured by the U-Prevent logbook. Higher scores indicate higher fidelity and adherence to the study protocol. Penetration is calculated as the number of healthcare professionals who deliver the PROSPERA program on an individual patient level, divided by the number of healthcare professionals who are expected to deliver this service. Implementation outcomes will be classified according to the taxonomy as proposed by Proctor et al. (29).

In this pragmatic cRCT we aim to standardize the function and the interconnected components of the PROSPERA program but acknowledge the intricacy of a complex intervention and how the components relate to each other (14). Therefore, adaptation to the context of a specific primary care practice is not directly seen as a deviation from the intervention as long as the general principle of the PROSPERA program, namely providing proactive and tailored care to meet the true needs of an individual patient, was not lost in the process (30).

#### Healthcare professional outcomes

Healthcare professionals’ satisfaction on usability of the PROSPERA program will be measured using the validated System Usability Scale (SUS), where higher scores indicate higher usability (31).

#### Patient outcomes

Perceived shared decision making and decisional conflict will be measured in a subgroup of 15% of patients, randomly selected by the healthcare professional. Shared decision-making will be assessed using the validated 9-item Shared Decision Making Questionnaire (SDM-Q-9) for patients (32). Higher scores indicate a greater extent of shared decision-making. Decisional conflict will be assessed using the validated Decisional Conflict Scale (DCS) for low literacy (33). Higher scores indicate more decisional conflict.

#### Cost effectiveness

The cost effectiveness of the PROSPERA program in primary care practices in the Netherlands will be assessed using a cost-utility analysis with Quality Adjusted Life Years (QALYs) and life-years (LYs) as the two primary effectiveness measures. A lifetime time horizon and a societal perspective will be applied. The trial results will be combined with other data sources to estimate the long-term costs and effects of PROSPERA. Besides multiple one-way sensitivity, subgroup and scenario analyses, a probabilistic sensitivity analysis will be conducted to quantify the uncertainty surrounding the incremental costs, effectiveness and incremental cost-effectiveness ratio (ICER). A cost-effectiveness acceptability curve will show the likelihood that PROSPERA is cost effective at different willingness-to-pay (WTP) thresholds. Lastly, a budget impact analysis (BIA) for implementation of the PROSPERA program in the Netherlands (minimum period, 3 years) will be performed based on data collected via ELAN and other sources. Both the CEA and BIA will be performed according to the Dutch guidelines for economic evaluations in healthcare (34).

### Sample size calculation

Since all enrolled patients will receive the population-level intervention component (empanelment), but only a subgroup will receive the additional individual patient level components, we have pre-specified a subanalysis of the primary outcome in this subgroup receiving all components. Since the clinical and implementation importance of this subanalysis, we chose to determine the sample size based on the required minimum number of patients expected to complete all components of the PROSPERA program. The study is powered to find a clinically relevant difference of 10% between the care-as-usual subgroup and the PROSPERA subgroup in reaching all protocol-defined cardiovascular preventive treatment goals. In the Netherlands, the standard practice size per full time equivalent GP is 2095 patients (35). On average, two to three GPs work together in a single primary care practice. Based on insights from literature and findings from our previously conducted qualitative study, we conservatively estimated that approximately 350 patients per 1 full time equivalent GP will meet the inclusion criteria (4). Consulted healthcare professionals indicated that a number of 40 patients per primary care practice will be feasible to treat following all components of the PROSPERA program during the course of the study. A power of 80% and an alpha of 5% were chosen as the statistical parameters for the sample size rationale. Assuming an intraclass correlation coefficient of 0.05 based on previous literature (36-38), this resulted in an estimated sample size of 11 clusters (primary care practices) with 35 patients per cluster and a total of 385 patients per study arm. This number of patients is in line with what healthcare professionals expected to be feasible. Including a loss-to-follow-up rate of 10%, the total sample size amounts to 848 patients over 22 primary care practices, with 424 patients in the intervention arm receiving the individual patient-level components of the PROSPERA program.

### Statistical analysis

The primary outcome will be analysed following the intention-to-treat principle. A mixed model is developed to determine the proportion of patients who have achieved all protocol-defined cardiovascular preventive treatment goals in the intervention group compared to the control group at the end of the study versus at the start. The start of the study was defined as the moment of PROSPERA empanelment. Data to assess the primary outcome will be extracted from ELAN using a dataset with an extraction date closest after the end of the follow-up period. Bias from measurement errors is expected to be randomly distributed across intervention and control practices since routine healthcare data are used to assess the outcomes of this study. Categorical data will be presented as numbers and percentages. Continuous variables will be summarized using means and standard deviations for approximately normally distributed data, and medians with interquartile ranges for skewed distributions.

In addition to the primary population analysis, we will perform a pre-specified subanalysis across all included practices for patients in the intervention group who received all components of the PROSPERA program. Further subgroup analyses will be performed for age and sex. We will also conduct a within patient comparison between pre- and post-PROSPERA program for included patients who received all components of the PROSPERA program. These outcomes will be analysed using multivariable logistic regression, adjusted for confounders, with GP practice included as random effect to account for clustering of patients within practices. A p-value of 5% will be considered clinically significant.

Since missing data on relevant CVD prevention treatment goals may move participating patients into a higher panel, we anticipate that this could partially affect the primary outcome. To account for this, a sensitivity analysis will be performed on multiple imputed treatment indicators.

The implementation outcomes, fidelity and penetration, and the questionnaires will be analysed using descriptive statistics. For the assessment of fidelity, we will utilise user data on adherence to the U-Prevent component of the PROSPERA program. Categorical data will be presented as numbers and percentages. Continuous variables will be presented as means with standard deviations or median and interquartile range, as appropriate.

#### Exploratory analyses

In our exploratory analyses we will also examine the difference between the intervention- and control group in 1) the net improvement of PROSPERA panel distribution, 2) the number of patients (%) that remained stable in the normal risk panel over the course of the study, 3) the improvement in other clinical patient related outcomes between pre- and post-implementation of the intervention between baseline and follow-up, i.e. in terms of risk score (%), LDL (absolute reduction in mmol/l) and systolic blood pressure (absolute reduction in mmHg), 4) the proportion (%) of patients with complete risk factor data registered between baseline and end of follow-up, 5) the absolute number (or %) of patients with (re)admission to the cardiovascular or diabetes mellitus integrated care programs between baseline and end of follow-up and 6) the difference between high- and low-socioeconomic position practices in reaching the primary outcome.

### Clinical implications

The PROSPERA program may have an impact on both the primary care practice and the individual patient level. At the level of primary care practices, a standardized risk-based structure for prioritized outreach and subsequent follow-up of the population of patients at risk of CVD is currently absent in the Netherlands. Primary care practices use a wide variety of approaches and invite patients opportunistically or reactively, based on surname, date of birth or the date of their previous visit. In a healthcare climate where scarcity is becoming ever more prominent, the PROSPERA program proposes an alternative proactive risk-based population approach for patients at risk of CVD in primary care. We expect the program to provide support to general practitioners and practice nurses in prioritizing high-risk cardiovascular patients, i.e., the patients who need urgent attention or intensification of preventive care. Simultaneously, it has the potential to provide a structured approach to reducing follow-up frequency for low-risk cardiovascular patients, who could instead be supported by remote patient monitoring or increased self-management. Insights from this study regarding the proactive population approach to CVRM of the PROSPERA program could be extended to other countries in which primary care is structured similarly with the GP as gatekeeper, such as the United Kingdom, Canada, Australia, New Zealand, and Scandinavian countries. However, since complex interventions such as the PROSPERA program are context-dependent, we assume that in countries where a risk-based outreach is already in place, the potential for achieving additional impact in terms of equity becomes smaller, and research into factors that hinder or facilitate sustainable implementation should be conducted.

At the individual patient level, the PROSPERA program is expected to result in improvement of traditional clinical risk predictors such as a decrease in LDL-cholesterol or systolic blood pressure through intensive counselling as part of the PROSPERA program, with a focus on tailored prevention and a basis in enhanced risk communication in combination with the visualizations of the patients’ individual treatment effect and cardiovascular-free life expectancy in U-Prevent. Similarly, we expect the visualizations and the Lifestylecheck to contribute to the process of shared decision making and decrease in decisional conflict (13). Therefore, even without a statistically significant improvement in achievement of traditional clinical risk factors in individual patients, the current trial may lead to valuable insights in how the different components of PROSPERA may interact. These insights may explain how PROSPERA might be meaningful in improving behavioural- and lifestyle factors through the emphasis on internal- and external motivation of a patient, personal drive and the increased attention to contextual factors that play a role in lifestyle change or treatment adherence necessary for the prevention of CVD. According to recent guidance of the MRC, such insights in inner mechanics of complex interventions are necessary for the improvement of development- and implementation of such interventions (14).

### Patient involvement

During the design of the study and writing of the protocol, a patient representative of Harteraad, a patient advisory council for heart disease, was involved.

## Supporting information

Supplemental Material Empanelment

Supplemental Material PROSPERA Fidelity Scale

## Data Availability

All data produced in the present study are available upon reasonable request to the authors.

## ETHICAL CONSIDERATIONS AND DISSEMINATION

This study will be conducted according to the principles of the Declaration of Helsinki (October 2024) and in accordance with the General Data Protection Regulation (version OJ L 119, 04.05.2016; cor. OJ L 127, 23.5.2018). The Medical Ethical Review Board of Leiden University Medical Center mandated the review of research not falling under the Dutch Medical Research with Human Subjects Law (nWMO) to individual nWMO committees at Leiden University Medical Center. The nWMO committee reviewed the study proposal and provided a declaration of no objection (non-WMO approval number: 23-3050). This study has been further approved by the scientific committee of the department of Public Health and Primary Care at Leiden University Medical Center.

Consent for the use of routine healthcare data was obtained through an opt-out procedure for participating practices. For the study questionnaires listed in table 1 we will obtain digital informed consent via Castor EDC (17). The trial is registered at http://www.clinicaltrials.gov under registration number NCT06593704. Results on effectivity, efficacy and implementation will be disseminated via peer reviewed journals and via (inter)national conferences to the scientific community and the public. Participating primary care practices and other stakeholders will receive update newsletters during the study about its progression and will be informed per email about the results. Results are expected in 2028.

## Acknowledgements

We thank our partners who contributed to the study: Harteraad, HADOKS, Stichting Langerhans, Stichting Informatievoorziening voor Zorg en Onderzoek (STIZON), Instituut voor Zorgoptimalisatie (INSZO) and ORTEC B.V. We especially thank H. J. de Jong for his help with the ELAN data extractions and data transfers.

## Contributors

VB, HO and SH have written the protocol of the study. MN, RV and JD substantially revised the protocol. VB wrote the draft of the manuscript. WR has written the section on cost-effectiveness and analyses. All authors read and substantially revised all versions of the manuscript. All authors gave final approval of the published version.

## Patient and public involvement

Patient advisory organisations were involved in the design and conduct of this research. Refer to the Methods section for further details.

## Patient consent for publication

Not applicable

## Provenance and peer review

Not commissioned

## Open access

This is an open access article distributed in accordance with the Creative Commons Attribution 4.0 Unported (CC BY 4.0) license, which permits others to copy, redistribute, remix, transform and build upon this work for any purpose, provided the original work is properly cited, a link to the licence is given, and indication of whether changes were made. See: licenses/by/4.0/.

