## Supplemental Material Empanelment for "Improving cardiovascular population risk in primary care: protocol for the PROSPERA cluster-randomized controlled trial of a complex multilevel intervention"

### Appendix A – Empanelment

**Explanatory notes for figure 1, table 1 and table 2 in this appendix**

The PROSPERA empanelment diagram (figure 1) is classified for cardiovascular risk and attainment of preventive cardiovascular treatment goals only. Care complexity arising from other factors (comorbidity, psychosocial complexity, etc.) is outside the scope of this project.

Protocol defined treatment goals in the PROSPERA study are in line with the Step 1 treatment targets as established by the 2021 European Society of Cardiology (ESC) Guidelines on cardiovascular disease prevention in clinical practice, the Dutch Cardiovascular Risk Management Guidelines of General Practitioners and the Dutch Diabetes Mellitus type 2 Guidelines of General Practitioners.

The achievement or non-achievement of lifestyle goals does not count towards the classification in the empanelment diagram. However, not achieving other applicable step 1 treatment targets, does result in additional support for achieving lifestyle goals by the primary care healthcare professional.

Apparently healthy patients are the patients requiring primary cardiovascular prevention. They have not yet experienced CVD.

Patients with a history of CVD are the patients who require secondary cardiovascular prevention.

We acknowledge that age not always accounts for vitality or vulnerability and that numerical treatment goals represent *target* values.

**Figure 1**. Empanelment diagram

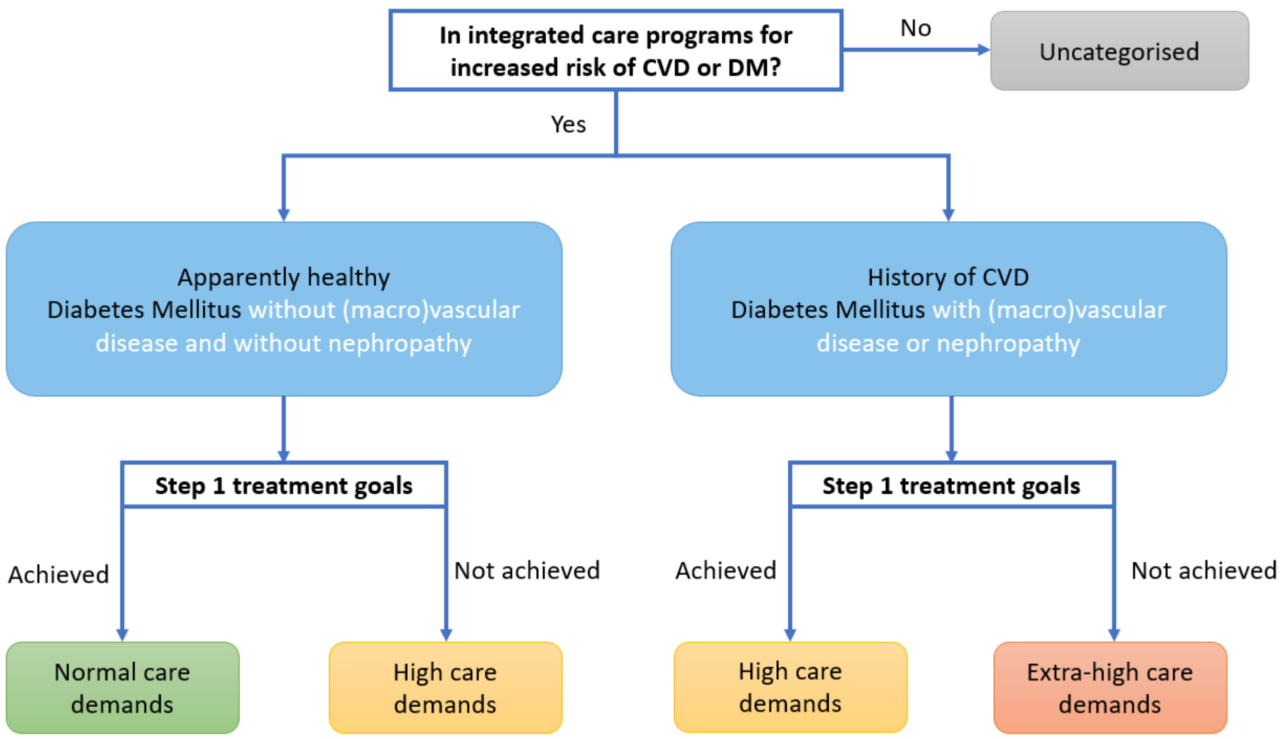

**Table 1.** Empanelment classification

| PROSPERA panels | Classification |
|---|---|
| Normal | • Apparently healthy, all step 1 treatment goals are reached<br>• Diabetes mellitus, all step 1 treatment goals are reached |
| High | • Established ASCVD*, all step 1 treatment goals are reached<br>• Apparently healthy, **not** all step 1 treatment goals are reached<br>• Diabetes mellitus **without** established ASCVD or nephropathy, **not** all step 1 treatment goals are reached |
| Extra high | • Established ASCVD, **not** all step 1 treatment goals are reached<br>• Diabetes mellitus **with** established ASCVD or nephropathy, **not** all step 1 treatment goals are reached |

**ASCVD = Atherosclerotic Cardiovascular Disease*

**Table 2.** Protocol defined treatment goals

| Apparently healthy + type of disease | Step 1 treatment goals |
|---|---|
| Age <70 years | Lifestyle*<br>Systolic blood pressure <140 mmHg<br>LDL-cholesterol <2.6 mmol/L |
| Age ≥70 years | Lifestyle<br>Systolic blood pressure <150 mmHg |
| + ASCVD (<70 years of age) | Antithrombotic therapy<br>LDL-cholesterol < 1.8 mmol/L |
| + ASCVD (≥70 years of age) | Antithrombotic therapy |
| + Diabetes Mellitus <70 years of age | Hba1c <53 mmol/mol |
| + Diabetes Mellitus ≥70 years of age | Hba1c <64 mmol/mol |
| + Diabetes and nephropathy | SGLT2i** |
| + Diabetes and arterial disease | SGLT2i and/or GLP1-RA*** |

**Lifestyle goals do not account for the classification but are considered step 1 treatment goals and treated as such. **Sodium-Glucose Linked Transporter 2 inhibitor. ***Glucagon-Like Peptide-1 Receptor Agonist.*
