## Supplemental Material PROSPERA Fidelity Scale for "Improving cardiovascular population risk in primary care: protocol for the PROSPERA cluster-randomized controlled trial of a complex multilevel intervention"

### Appendix B – PROSPERA Fidelity Scale

PROSPERA Fidelity Scale – Part 1 (Month 4)

**Introduction**
This questionnaire contains questions regarding the use of the PROSPERA program in your primary care practice thus far.

**Part A. General**

**Question 1. Year of birth**
What is your year of birth?

*[numeric entry]*

**Question 2. Gender**
What is your gender?

*[option group]*

- *Female*
- *Male*
- *Other*

**Question 3. Professional role**
What is your professional role?

*[option group]*

- *Practice nurse (PN)*
- *Nurse specialist (NS)*
- *General practitioner (GP)*
- *GP in training*

**Question 4. Workplace**
Where do you work?

*[name of general practice]*

**Question 5. Work experience**
How many years of work experience do you have as a PN, NS, GP, etc.?

*[numeric entry]*

**Part B. Training and start**
The following questions concern the training and the start of the study.

**Question 6. Training**
Did you attend the PROSPERA training on 09-10-24?

*[option group]*

- *Yes*
- *No*

**Question 7. Recording**
Did you watch the recording of the PROSPERA training on 09-10-24 online afterwards?

*[option group]*

- *Yes*
- *No*

**Question 8. Kick-off visit**
Were you present at the kick-off visit?

*[option group]*

- *Yes → if yes, then question 8.1*
- *No*

**Question 8.1. Kick-off visit_month**
In which month did the kick-off visit take place at the practice?

*[option group]*

- *October 2024*
- *November 2024*
- *December 2024*
- *January 2025*
- *February 2025*
- *March 2025*
- *This took place later than March 2025*
- *This never took place*

**Information**
In addition to the training, the PROSPERA program consists of several other components:

1. Proactive selection and invitation
2. Use of the Lifestylecheck
3. Use of the automatically pre-filled U-Prevent application

The following questions concern these components of the PROSPERA program.

**Question 9. Intention**
Before the study started, did you intend to work with all components of the PROSPERA program?

*[option group]*

- *Yes*
- *No → if no, then question 9.1*

**Question 9.1. Fidelity_prior**
Which component of the PROSPERA program did you expect beforehand to use less or differently than intended?

*[option group, multiple answers possible]*

- *Proactive selection and invitation*
- *The Lifestylecheck*
- *The automatically pre-filled U-Prevent application*

**Question 9.2.** Explanation? *(Optional)*
..........................................................................................................................................
..........................................................................................................................................
..........................................................................................................................................
.......................................................

**Question 10. Completeness**
Have you worked with <u>all components</u> of the PROSPERA program thus far?

*[option group]*

- *Always*
- *Almost always*
- *Sometimes*
- *Almost never*
- *Never*

**Question 10.1.** Explanation? *(Optional)*
..........................................................................................................................................
..........................................................................................................................................
..........................................................................................................................................
.......................................................

**Question 11. SOP**
How often have you consulted the manual <u>thus far</u>?

*[option group]*

- *Never*
- *1–2 times*
- *3–4 times*
- *5–10 times*
- *More than 10 times*

**Information**
The following questions concern the individual components of the PROSPERA program. Indicate how often you have used specific components of the PROSPERA program <u>thus far</u>.

**Question 12. Proactive invitation**
Have you thus far invited patients with an increased cardiovascular risk (including patients with diabetes) <u>with prioritization</u>? *Check all that apply.*

*[option group]*

- *Always*

- *Almost always*
- *Sometimes*
- *Almost never*
- *Never*

**Question 12.1.** Explanation? (*Optional*)
……………………………………………………………………………………………………………………………………
……………………………………………………………………………………………………………………………………
……………………………………………………………………………………………………………………………………
……………………………………………………

**Question 13. Shared decision-making**
How often have you thus far conducted a shared decision-making consultation with patients using **both** components mentioned above **(Lifestylecheck and U-Prevent**)?

*[option group]*

- *Always*
- *Almost always*
- *Sometimes*
- *Almost never*
- *Never*

**Question 13.1.** Explanation? (*Optional*)
……………………………………………………………………………………………………………………………………
……………………………………………………………………………………………………………………………………
……………………………………………………………………………………………………………………………………
……………………………………………………

**Question 14.**
On a scale from 0 (never) to 100 (always): how often have you thus far conducted a shared decision-making consultation with patients using **only the Lifestylecheck**?

0 ——————————————————————— 100

**Question 15.**
On a scale from 0 (never) to 100 (always): how often have you thus far conducted a shared decision-making consultation with patients using **only the automated version of U-Prevent**?

0 ——————————————————————— 100

**Question 16.** Explanation? (*Optional*)
……………………………………………………………………………………………………………………………………
……………………………………………………………………………………………………………………………………
……………………………………………………………………………………………………………………………………
……………………………………………………

**Closing**
This was the final question of this questionnaire. Thank you very much for completing it!

<u>PROSPERA Fidelity Scale – Part 2 (Month 10)</u>

**Information**
In addition to the training, the PROSPERA program consists of the following components:

- Proactive selection and invitation
- Use of the Lifestylecheck
- Use of the automatically pre-filled U-Prevent application

**Question 1. Completeness**
Have you thus far worked with all components of the PROSPERA program?

*[option group]*

- *Always*
- *Almost always*
- *Sometimes*
- *Almost never*
- *Never*

**Question 1.1.** Explanation? (*Optional*)
.........................................................................................................................................
.........................................................................................................................................
.........................................................................................................................................

**Question 2. SOP**
How often have you consulted the user manual thus far?

*[option group]*

- *Never*
- *1–2 times*
- *3–4 times*
- *5–10 times*
- *More than 10 times*

**Question 3. Proactive invitation**
Have you thus far invited patients with an increased cardiovascular risk (including patients with diabetes) <u>with prioritization</u>? *Check all that apply.*

*[option group]*

- *Always*
- *Almost always*
- *Sometimes*
- *Almost never*
- *Never*

**Question 3.1.** Explanation? (*Optional*)
.........................................................................................................................................

……………………………………………………………………………………………………………………
……………………………………………………………………………………………………………………

**Question 4. Shared decision-making**
How often have you thus far conducted a shared decision-making consultation with patients using **both** the Lifestylecheck and U-Prevent?

*[option group]*

- *Always*
- *Almost always*
- *Sometimes*
- *Almost never*
- *Never*

**Question 4.1.** Explanation? (*Optional*)
……………………………………………………………………………………………………………………
……………………………………………………………………………………………………………………
……………………………………………………………………………………………………………………

**Question 5.**
On a scale from 0 (never) to 100 (always): how often have you conducted a shared decision-making consultation with patients using **only the Lifestylecheck**?

0 ———————————————————————————— 100

**Question 6.**
On a scale from 0 (never) to 100 (always): how often have you conducted a shared decision-making consultation with patients using **only the automated version of U-Prevent**?

0 ———————————————————————————— 100

**Question 7. Fidelity_during**
Was there any component of the PROSPERA program that you used differently than intended? *Examples include completing the Lifestylecheck during the consultation instead of beforehand or switching to the non-automated version of U-Prevent.*

*[option group]*

- *Yes*
- *No*

**Question 7.1. Which component did you use differently than intended?**
*[option group, multiple answers possible]*

- *Proactive selection and invitation*
- *The Lifestylecheck*
- *The automatically pre-filled U-Prevent application*

**Question 7.2.** Explanation? *How did you use it differently?*
……………………………………………………………………………………………………………………

..................................................................................................................................
..................................................................................................................................

**Question 8. Target population**
On a scale from 0 (never) to 100 (always): were you able to use the PROSPERA program for all cardiovascular disease (CVD) patients you had selected? In what percentage of cases do you think this was successful?

0 ———————————————————————————— 100

**Question 9.** Explanation? (*Optional*)
..................................................................................................................................
..................................................................................................................................
..................................................................................................................................

**Question 10. Early discontinuation**
The intention was to use the PROSPERA program for 10 months. Did you discontinue the program prematurely?

*[option group]*

- *Yes*
- *No*

**Question 10.1. Duration of the program**
For how long were you able to use the PROSPERA program?

*[option group]*

- *I used it for 7 to 10 months*
- *I used it for 3 to 6 months*
- *I used it for 0 to 3 months*
- *We never started*

**Question 10.2.** Explanation? (*Optional*)
..................................................................................................................................
..................................................................................................................................
..................................................................................................................................

**Question 11.**
Do you have any additional comments or anything else you would like to add?

**Closing**
This was the final question of this questionnaire. Thank you very much for completing it!
